# Heterogeneity in non-suicidal self-injury ideation and treatment response among inpatient female adolescents with mood disorders: Insights from ecological momentary assessment

**DOI:** 10.64898/2026.09.16.26362947

**Authors:** Meiping Zeng, Yanhuan Huang, Yue Ding, Zhishan Hu, Zhen Liu, Changminghao Ma, Wenjing Liu, Xifeng Xia, Jiayi Song, Fang Zhang, Wenhong Cheng

## Abstract

Non-suicidal self-injury (NSSI) is highly prevalent and exhibits substantial fluctuations among adolescents with mood disorders, yet the heterogeneity and differential treatment responses accompanied by such fluctuations remain unclear. The current study employed ecological momentary assessment (EMA) and latent profile analysis to identify latent subtypes of NSSI ideation via temporal features, and constructed machine-learning models to validate the predictive value of dynamic features for post-treatment self-injury risk. Thirty-five Chinese female adolescents with mood disorders and co-occurring NSSI (*M*_age_ = 14.4 years) receiving inpatient psychiatric treatment completed baseline assessments and 5 daily surveys of NSSI ideation throughout the course of treatment. Two subtypes were identified: the high-mean paroxysmal subtype (54.3%) and the low-mean sporadic subtype (45.7%). The high-mean paroxysmal subtype exhibited significantly more severe clinical symptoms, more emotional abuse experiences, lower self-esteem, higher impulsivity and greater difficulties in emotion regulation. The temporal-feature-based model outperformed the baseline-feature-based model in predicting self-injury risk. These findings underscore the dynamic nature and heterogeneity of NSSI ideation among adolescents with mood disorders, and offering implications for tailoring interventions to distinct youth subgroups.

## 1 Introduction

Non-suicidal self-injury (NSSI) refers to direct and deliberate damaging of one’s body issue without suicidal intent, and such behaviors are generally not culturally sanctioned (Nock, 2010). Meta-analysis has reported a pooled prevalence of 17.7% for NSSI among adolescents aged 10-19 years (Denton & Álvarez, 2024). In China, the pooled prevalence ranges from 25.3% to 32.8% (Qu et al., 2023), and it rises as high as 54% among adolescents with mood disorders seeking psychiatric care (Yang et al., 2022). These findings highlight the clinical importance of NSSI, a condition formally recognized by the American Psychiatric Association in the Diagnostic and Statistical Manual of Mental Disorders, Fifth Edition, Text Revision (DSM-5-TR; APA, 2022). Notably, the prevalence of NSSI is significantly higher in female than male adolescents (26.5% vs. 13.7%; Denton & Álvarez, 2024), and this gender difference remains evident among adolescent patients with mood disorder (Liu et al., 2023). NSSI not only serves as a robust predictor of future suicidal attempts (Daukantaitė et al., 2021), but is also associated with high comorbidity rates across multiple mental disorders, especially depressive disorders, anxiety disorders, and borderline personality disorder (Chen et al., 2025). Despite progress in understanding the prevalence, correlates, and longer-term course of NSSI among individuals seeking treatment, little is known about its short-term course across days and weeks during treatment. Consequently, effective intervention remains challenging.

Despite the shared clinical features of NSSI, growing evidence suggests considerable heterogeneity among adolescents engaging in self-injurious behaviors (Coppersmith et al., 2021;Wu et al., 2022). Compared with traditional classification approach, person-centered approaches, such as latent profile analysis (LPA), can accurately capture heterogeneous profiles based on observed frequency, severity and other factors (Demaray et al., 2021). A systematic review indicated that, based on NSSI characteristics such as frequency and function, individuals with NSSI can typically be classified into two to five subtypes (Fox et al., 2020). For instance, de Neve-Enthoven et al. (2024) identified four subtypes of high-risk adolescents with distinct combinations of NSSI and suicide levels (low/moderate/high). Similarly, He et al. (2023) distinguished two subtypes, including high and low suicidal ideation, among Chinese adolescents with depression. In addition, several studies have differentiated subtypes according to psychosocial traits, consistently showing that high-risk NSSI subtypes are significantly associated with severe clinical symptoms, low self-esteem, and elevated impulsivity (Lee et al., 2024; Li et al., 2026). Collectively, these findings support the heterogeneity inherent in the NSSI population and highlights the need to further explore distinct subtypes to improve screening accuracy and intervention specificity.

Recommended by the National Institute for Health and Care Excellence (NICE, 2022), Dialectical Behavior Therapy for Adolescents (DBT-A) is a first-line psychological intervention for adolescents with recurrent self-harm. Through structured skills training, such as mindfulness, distress tolerance, and emotion regulation, DBT-A aims to equip adolescents with healthy coping strategies that serve as alternatives to self-harm (Miller et al., 2007; Rathus & Miller, 2015). Although DBT-A is supported by robust empirical evidence (Kothgassner et al., 2021), its cost-effectiveness poses significant challenges. An economic analysis by Mavranezouli et al. (2024) reported that the incremental cost-effectiveness ratio of DBT-A compared with enhanced usual care was £268,601 per quality-adjusted life year (QALY), exceeding the threshold recommended by NICE in the UK. Limited by its long duration (6-12 months), high treatment demands, and shortages of specialized therapists, many eligible adolescents remain unable to access DBT-A (White et al., 2023). In addition, Berk et al. (2022) found that approximately 13% of DBT-A recipients exhibited non-response to treatment with increasing self-injury from mid-treatment to follow-up. Such poor outcomes may be significantly associated with higher baseline pessimism, depressive symptoms, and perceived burden (Abbott et al., 2019). These findings indicate that solely relying on group-level averages may obscure important individual differences in treatment response. Accordingly, further characterize heterogeneity in NSSI population is warranted to better identify adolescents most in need of prioritized DBT-A.

Notably, self-injurious ideation and behaviors are short-lived and highly fluctuating (Coppersmith et al., 2021; Fitzpatrick et al., 2020). These features are difficult to capture in prior subtyping studies relying on cross-sectional or retrospective designs with coarse temporal resolution (e.g., He et al., 2023; Reinhardt et al., 2022). In contrast, ecological momentary assessment (EMA), which collects intensive real-time data on individuals’ momentary states through repeated measurements over a short period, can effectively capture the dynamic shifts in psychological processes (Hoelscher et al., 2025). EMA has demonstrated high feasibility and adherence in in adolescent NSSI research, with meta-analytic evidence showing high compliance in daily-life assessments (Martin et al., 2026) and no evidence of elevated self-harm or suicide risk from repeated measurement (Cycz et al., 2018). Early research found that most self-injurious thoughts among adolescents in daily life are brief (i.e. 1-30 minutes), moderate in intensity, and typically precede self-injurious behaviors (Nock et al., 2009). Compared with retrospective interviews, adolescents report more self-injurious events via EMA (Esposito et al., 2022). Furthermore, in a 28-day EMA study of 125 adolescents, Kiekens et al. (2024) found that NSSI ideation and urges fluctuate considerably both between and within individuals, with over one-fifth of participants showing meaningful changes across assessments spaced less than two hours apart. Taken together, EMA holds substantial value for capturing the dynamic fluctuations and individual differences of NSSI ideation.

However, in existing self-harm and suicide research, EMA has been predominantly used to monitor within-person variability in self-injurious thoughts or to explore time-varying associations between antecedent variables and NSSI behaviors (Huang et al., 2025). Few studies have directly adopted EMA to subtype NSSI ideation, especially in clinical adolescents. In clinical adult samples, Kleiman et al. (2017) conducted an EMA study with 32 adults and identified five subtypes of suicidal ideation via temporal features (i.e., intensity, frequency, and variability). This finding was later replicated in 27 Chinese adult inpatients (Wu et al., 2022). Nonetheless, EMA research focusing on NSSI subtypes in clinical adolescent samples remains scarce, which further limits the exploration of associations between dynamic changes in self-injury ideation and differential treatment responses. Moreover, the onset and maintenance of NSSI involve a range of vulnerability and precipitating factors (Nock, 2009), for example, childhood trauma, low self-esteem, emotion dysregulation, and impulsivity (Qu et al., 2023). In this context, traditional regression approaches are unsuited to capture interactions among diverse types of predictors, thereby limiting their accuracy in predicting treatment responses. In recent years, machine learning approaches have been applied to predict suicide-related behaviors in adolescents, with meta-analytic evidence supporting their high specificity (Liu et al., 2025). In a pioneering study, Wang et al. (2021) combined EMA with machine learning in adult inpatients, and demonstrated that temporal features derived from EMA exhibited superior performance for post-treatment suicide attempts compared with standardized baseline assessments. These findings suggest that intensive longitudinal monitoring may yield prognostic insights beyond static assessments, highlighting the need to integrate dynamic NSSI features with subtyping in adolescent clinical samples.

In summary, NSSI is highly prevalent and heterogeneous among adolescents with mood disorders. Existing subtype studies predominantly rely on retrospective assessments with coarse temporal granularity, which fail to capture the short-term fluctuating nature of NSSI ideation. Although EMA facilitates capturing dynamic psychological changes, research applying EMA to NSSI subtyping among clinical adolescents remains limited. Integrating machine learning with EMA may also provide novel insights into individual differences in treatment response. Therefore, the current study first aims to leverage intensive EMA data from inpatient female adolescents with NSSI to identify latent subtypes of NSSI ideation and characterize their clinical characteristics and intervention outcomes. In addition, we also construct support vector machine (SVM) models to validate the predictive utility of temporal temporal features for treatment response, so as to provide empirical references for the application of intensive dynamic monitoring in precision-oriented clinical intervention.

## 2 Methods

### 2.1 Participants

All the participants were recruited from the inpatient wards of the Shanghai Mental Health Center from March to August 2024. The inclusion criteria were (1) age between 12 and 18 years, (2) meeting DSM-5 diagnostic criteria for NSSI, and (3) past-week high self-injury risk, defined as a score of 6 or higher on the Alexian Brothers Urge to Self-injure Scale (Bahamon et al., 2023).

Exclusion criteria included: (1) bipolar disorder, substance and alcohol abuse, autism spectrum disorder, intelligence disorder, and schizophrenia that are unable to comprehend or independently complete the EMA protocol; (2) refused to receive the routine inpatient psychotherapy; (3) completed less than 10 valid EMA responses within the first week of admission. Diagnosis and eligibility were determined by an experienced chief psychiatrist through structured clinical interviews and medical record reviews. All participants and their parents provided written informed consent. This study was approved by the Ethics Committee of Shanghai Mental Health Center (NO. 2022-74C2).

A total of 45 adolescents were initially enrolled, of whom 10 were excluded after professional evaluation for not meeting the inclusion criteria. The final sample consisted of 35 female adolescents, with a mean baseline age of 14.4 years (*SD* = 2.2). Although the sample size is modest, this study adopted an intensive longitudinal EMA design with repeated assessments within individuals. In EMA studies, statistical power for within-person analyses is determined by the number of observations per person (and thus the total observation count), rather than the number of participants (Bolger & Laurenceau, 2013). On average, each participant contributed 149 assessments, yielding a total of 5,242 momentary observations for analysis. It is also worth noting that all participants were female, though imbalanced, is broadly broadly representative of the sex distribution typically observed in adolescent inpatient psychiatric settings (Shi & Qiu, 2024). Taken together, the current sampling density was considered adequate to address the heterogeneity of NSSI ideation and its within-person associations.

### 2.2 Measurements

#### Baseline measurements

Demographic characteristics obtained, including sex, age and only-child status. Primary diagnoses and comorbidity were assesses using the Mini International Neuropsychiatric Interview for Children and Adolescents (MINI-Kids) and supplemented by medical record reviews.

The Chinese version of the Ottawa Self-Injury Inventory (OSI; Zhang et al., 2015) is a 32-item measure that provides a comprehensive assessment of non-suicidal self-injury behaviors. In the present study, selected items were administered to assess the frequency of NSSI over the past week, month, and year on a 4-point scale (0 = never, 3 = daily), along with age of onset, number of NSSI methods, and NSSI functions.

In terms of psychological characteristics, the Chinese version of Childhood Trauma Questionnaire (CTQ-SF; Zhang, 2011), Difficulties in Emotion Regulation Scale (DERS; Ding et al., 2013), Rosenberg Self-Esteem Scale (RSES; Wang et al., 1999), and Barratt Impulsiveness Scale (BIS; Li, Fei, Xu et al., 2011) were administered to assess childhood traumatic experiences, emotion regulation difficulties, self-esteem, and impulsivity traits, respectively. For the CTQ-SF, each of the five subscales yields scores ranging from 5 to 25. On the subscales, physical abuse ≥ 8, emotional abuse ≥ 9, sexual abuse ≥ 6, emotional neglect ≥ 10, and physical neglect ≥ 8 were used to define the presence of the corresponding types of child maltreatment (Pham et al., 2021).The BIS also comprises three dimensions: motor impulsivity, cognitive impulsivity, and non-planning impulsivity, with higher scores reflecting greater impulsivity in the corresponding domain.

Clinical symptoms, including depressive symptoms, anxiety symptoms, self-harm urge, and suicidal ideation, were assessed both at pre- and post-treatment using the Beck Depression Inventory (BDI-Ⅱ-C; Yang et al., 2024), the Screen for Child Anxiety Related Emotional Disorders (SCARED; Wang et al., 2002), the Alexian Brothers Urge to Self-Injure Scale (ABUSI; Washburn et al., 2010), and the Beck Suicidal Ideation - Chinese Version (BSI-CV; Li, Fei, Zhang et al., 2011), respectively. Higher total scores on each scale indicated more severe corresponding clinical symptoms. Notably, a total score of 6 or above on ABUSI was defined as high NSSI risk (Bahamon et al., 2023). Based on this cutoff, a binary variable (0 = low risk, 1 = high risk) representing post-treatment NSSI risk was generated for subsequent machine learning.

#### Ecological momentary assessment

Following the framework of Kleiman et al. (2017), three visual analog scale items were adopted during each EMA survey. Participants were asked to rate the momentary extent to NSSI urges (“Right now, how strong is the urge present to harm yourself without suicidal intent?”), NSSI intentions (“Since your last report, have you considered engaging in self-injury?”), and self-efficacy for resisting NSSI (“How confident are you that you will not engage in self–injury before your next report?”), respectively, rated from 0 (not at all) to 10 (extremely). After reverse-coding the self-efficacy item, the three items were summed to form a composite score of NSSI ideation, with higher scores indicating greater NSSI ideation.

Drawing on a systematic review of EMA studies among children and adolescents reporting an average sampling frequency of 4.4 assessments per day (Heron et al., 2017), as well as the high acceptability observed in our pilot test, the present study employed a five-times-daily EMA sampling protocol. During inpatient treatment, participants completed five daily EMA surveys on study-provided electronic devices between 8am and 8pm, with assessments semi-randomly assigned to five time windows: morning, noon, afternoon, evening, and before bedtime. Assessments were coded as missing when participants were unable to complete them due to concurrent therapy sessions. Participants received a small incentive (e.g. candy or stickers) each evening upon completion of daily assessments.

### 2.3 Data analysis

Analyses were conducted in R (Version 4.1.2) and IBM SPSS Statistics (Version 24). Missing data were handled using multiple imputations with the MICE package in R. For each participant, eight temporal features were respectively derived from EMA reports of NSSI urge, NSSI intensity, self-efficacy to resist NSSI, and NSSI ideation: mean, intra-individual standard deviation (SD), maximum (Max), non-zero proportion, root mean square of successive differences (RMSSD), percentage of unique values (PUV), probability of acute change (PAC), and maximum change (Max Change). The first four features describe the momentary intensity at the measurement occasions, whereas the latter four features reflect the dynamic variability across assessments.

Second, latent profile analysis (LPA) was undertaken to identify profiles based on the eight temporal features of NSSI ideation. The best fitting models were selected using a range of fit-statistics and substantive considerations. Lower values for information criteria such as Akaike Information Criterion (AIC), Bayesian Information Criterion (BIC), and Sample-size Adjusted Bayesian Information Criterion (aBIC), indicated better model fit (Si et al., 2022). Significant p-values of the Bootstrapped parametric Likelihood Ratio test (BLRT) indicated that an additional class could significantly improve the model. After profile enumeration, subtype differences were examined in terms of baseline NSSI, psychological characteristics, and clinical symptoms.

Third, two support vector machine (SVM) models were constructed to compare predictive performance for post-treatment self-injury risks. Prior to modeling, principal component analysis (PCA) was performed on each set to select the top-10 contributory baseline features and temporal features, respectively, ensuring an equal number of predictors across both models and reduced model complexity. The outcome variable was the binary classification (0 = low risk, 1 = high risk) representing post-treatment NSSI risk. Model performance was evaluated using accuracy, precision, specificity, sensitivity, and area under the ROC curve (AUC), with higher values indicating better predictive performance. For further details regarding mode-building procedures , see the supplementary materials.

## 3 Results

### 3.1 Descriptive statistics

The final sample consists of 35 female adolescents with ages ranging from 12 to 18 years and a mean age of 14.37 years (*SD* = 2.24). According to the MINI-KIDS and clinician review, 23 (65.71%) were diagnosed with depressive disorder, 8 (22.86%) with mood disorder, and 4(11.43%) with mixed anxiety and depressive disorder. Among these participants, 19 (54.29%) presented additional comorbidity. Regarding baseline NSSI, the mean age of NSSI onset was 11.77 years (*SD* = 1.73), and participants reported an average of 5 NSSI methods (*SD* = 2.41). A total of 27 participants (77.14%) had engaged in NSSI within the week prior to baseline assessment.

During the EMA data collection, participants were assessed for an average of 30.8 days (*SD* = 11.79), yielding a total of 5,242 EMA observations ( approximately 150 per participant). The overall compliance race was 98.03%, suggesting the positive attitude toward participation in the study among this sample of inpatient adolescents at risk of NSSI.

### 3.2 Latent profiles of NSSI ideation

LPA was performed using eight temporal features of NSSI ideation (i.e., mean, SD, Max, non-zero-proportion, RMSSD, PVA, PAC, Max change), and models with one to for profiles were estimated. Model fit indices for different latent profile models are presented in Table 1. From the one-profile to the three-profile solutions, AIC and aBIC decreased gradually and entropy increased, whereas BIC showed a slight increase. The four-profile solution yielded worse fit than the three-profile model due to the higher AIC, BIC and lower entropy. Moreover, the three-profile model produced a non-significant BLRT result (*p* = .050), and the additional profile in the three-profile solution only subdivided one of the existing profiles without revealing a distinct pattern. Thus, the two-profile solution was selected as the optimal model, with a minimum profile membership of 45.7%, ensuring adequate subgroup size for subsequent analyses.

**Table 1.** Model fit indices for LPA models representing one to four NSSI ideation groups.

| Classes | AIC | BIC | aBIC | BLRT($p$) | Entropy | Smallest<br>profile<br>size |
| --- | --- | --- | --- | --- | --- | --- |
| 1 | 908.60 | 977.03 | 839.64 | - | - | - |
| 2 | 897.09 | 979.52 | 814.03 | 0.040 | 0.972 | 45.7% |
| 3 | 889.36 | 985.79 | 792.20 | 0.050 | 0.987 | 22.9% |
| 4 | 899.98 | 1010.41 | 788.71 | 0.842 | 0.984 | 8.6% |

*Note.* AIC = Akaike Information Criterion, BIC = Bayesian Information Criterion, aBIC = Sample size-adjusted Bayesian Information Criterion, BLRT = Bootstrap Likelihood Ratio Test.

As shown in Figure 1, the following labels were assigned to the profiles: High-mean paroxysmal subtype (54.3%, *n* = 18, i.e., high NSSI ideation levels and marked fluctuations, with four change points observed at days 4, 12, 15, and 41 of the assessment period) and Low-mean sporadic subtype (45.7%, *n* = 17, i.e., low NSSI ideation levels and modest fluctuations, with a single change point at day 5 for the assessment period). Overall, most participants showed a decreasing trend in NSSI urges and intentions, alongside an increasing trend in self-efficacy to resist NSSI. Despite the similar numbers of peaks between the two subtypes, further comparisons on the eight temporal features of NSSI ideation revealed significant group differences across all features except maximum change (*p* = .126). Specifically, compared to the low-mean sporadic subtype, the high-mean paroxysmal subtype showed significantly higher values on mean (*p* < .001), SD (*p* < .001), Max (*p* = .001), non-zero proportion (*p* < .001), RMSSD (*p* < .001), and PUV (*p* < .001), but significantly lower values on PAC (*p* =.008). Differences between the two subtypes on the eight temporal features are illustrated in Figure 2.

**Figure 1.**
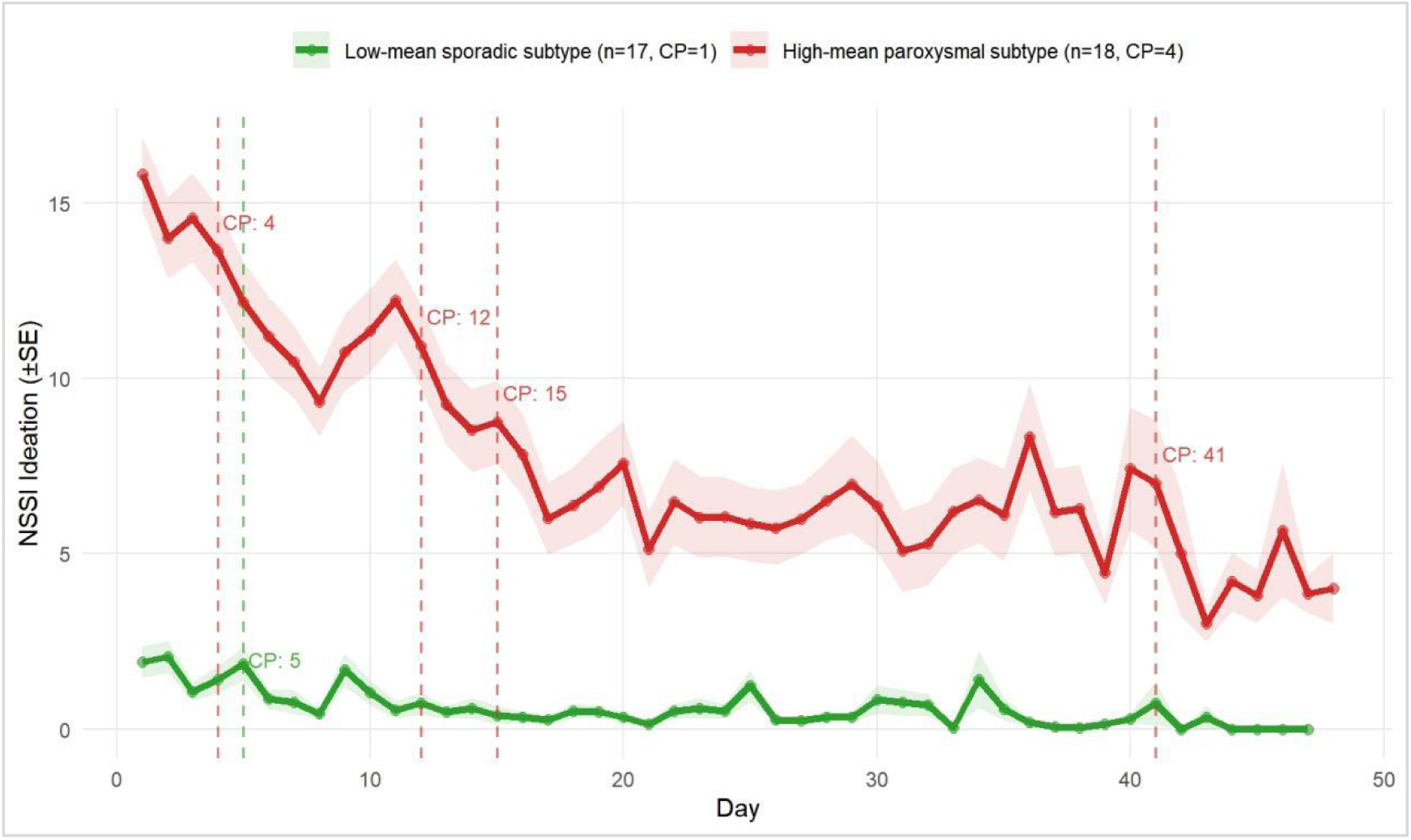
Subtype-based times series plot of NSSI ideation with change points

**Figure 2.**
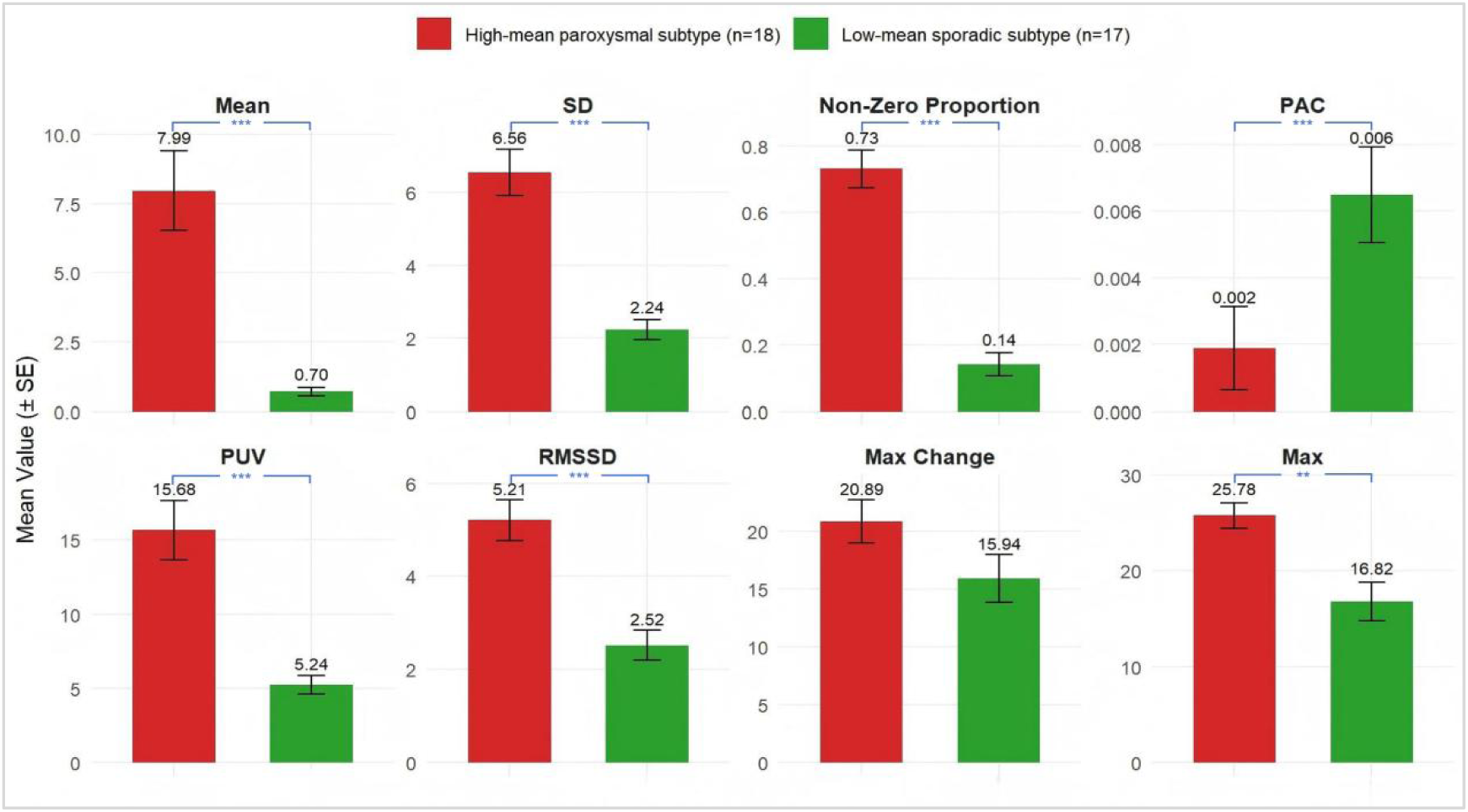
Comparisons between the two subtypes on the eight temporal features of NSSI ideation *Note.* SD = intra-individual standard deviation, PAC = probability of acute change, PUV = percentage of unique values, RMSSD =root mean square of successive differences, Max Change = maximum change. \*\**p*<.01, \*\*\**p*<.001.

### 3.3 Differences between subtypes of clinical symptoms and psychological characteristics

To further distinguish the two subtypes and identify the high-risk group, we compared the two subtypes on clinical symptoms and psychological characteristics (see Table 2). In terms of clinical symptoms, the high-mean paroxysmal subtype had significantly higher pre-treatment depressive symptoms (*U* = 84.00, *p* = .022) and self-harm urges (*t* = 2.11, *p* = .043) than the low-mean sporadic subtype, with no significant group differences on the other two clinical scales (*p* = .067; *p* = .163). Post-treatment, the high-mean paroxysmal subtype remained significantly higher on depressive symptoms (*U* = 85.00, *p* = .025), self-harm urges (*U* = 37.50, *p* < .001), and suicidal ideation (*U* = 69.50, *p* = .005), whereas anxiety symptoms did not differ significantly between groups (*p* = .053). Notably, the high-mean paroxysmal subtype’s post-treatment ABUSI total score remained above the clinical cutoff, and its BDI-II total score fell within the major depression range, suggesting poorer treatment outcomes for this subtype.

**Table 2.**
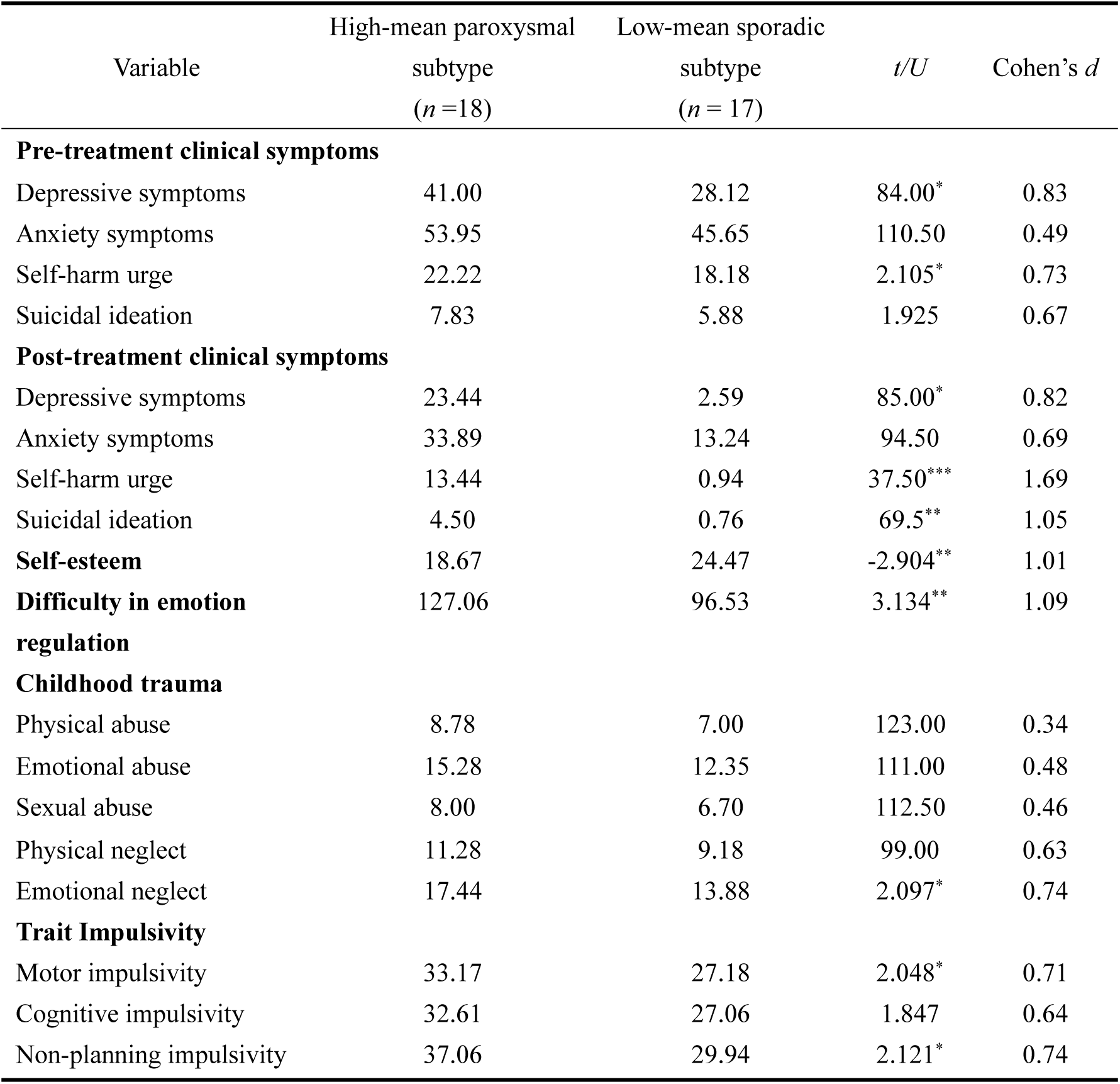
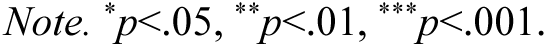
Differences in clinical symptoms and psychological characteristics across subtypes.

| Variable | High-mean paroxysmal<br>subtype<br>($n=18$) | Low-mean sporadic<br>subtype<br>($n = 17$) | $t/U$ | Cohen's $d$ |
| --- | --- | --- | --- | --- |
| **Pre-treatment clinical symptoms** |  |  |  |  |
| Depressive symptoms | 41.00 | 28.12 | 84.00* | 0.83 |
| Anxiety symptoms | 53.95 | 45.65 | 110.50 | 0.49 |
| Self-harm urge | 22.22 | 18.18 | 2.105* | 0.73 |
| Suicidal ideation | 7.83 | 5.88 | 1.925 | 0.67 |
| **Post-treatment clinical symptoms** |  |  |  |  |
| Depressive symptoms | 23.44 | 2.59 | 85.00* | 0.82 |
| Anxiety symptoms | 33.89 | 13.24 | 94.50 | 0.69 |
| Self-harm urge | 13.44 | 0.94 | 37.50*** | 1.69 |
| Suicidal ideation | 4.50 | 0.76 | 69.5** | 1.05 |
| **Self-esteem** | 18.67 | 24.47 | -2.904** | 1.01 |
| **Difficulty in emotion regulation** | 127.06 | 96.53 | 3.134** | 1.09 |
| **Childhood trauma** |  |  |  |  |
| Physical abuse | 8.78 | 7.00 | 123.00 | 0.34 |
| Emotional abuse | 15.28 | 12.35 | 111.00 | 0.48 |
| Sexual abuse | 8.00 | 6.70 | 112.50 | 0.46 |
| Physical neglect | 11.28 | 9.18 | 99.00 | 0.63 |
| Emotional neglect | 17.44 | 13.88 | 2.097* | 0.74 |
| **Trait Impulsivity** |  |  |  |  |
| Motor impulsivity | 33.17 | 27.18 | 2.048* | 0.71 |
| Cognitive impulsivity | 32.61 | 27.06 | 1.847 | 0.64 |
| Non-planning impulsivity | 37.06 | 29.94 | 2.121* | 0.74 |

*Note.* \* $p < .05$ , \*\* $p < .01$ , \*\*\* $p < .001$ .

Regarding psychological characteristics, adolescents in the high-mean paroxysmal subtype exhibited lower self-esteem (*t* = -2.90, *p* = .007), greater emotion regulation difficulties (*t* = 3.13, *p* = .004), and higher levels of emotional neglect experiences (*t* = 2.07, *p* = .044) compared with the low-mean sporadic subgroup. For impulsivity, the high-mean paroxysmal subtype also showed significantly higher motor impulsivity (*t* = 2.05, *p* = .049) and non-planning impulsivity (*t* = 2.12, *p* = .042).

### 3.4 Optimal model for predicting integrated treatment outcomes

Using principal component analysis, the top-10 contributory baseline features were as follows: three NSSI frequency (past-week, past-month, past-year); three NSSI functions (sensation-seeking, emotion-regulation, and interpersonal influence); baseline level of three clinical symptoms (anxiety symptoms, NSSI urges, and suicidal ideation); and only-child status. Similarly, derived from EMA data collected during the first week of admission, the top-10 contributory temporal features included three from momentary NSSI urges (mean, non-zero proportion, and RMSSD), three from momentary NSSI intentions (mean, non-zero proportion, and max change); and four from momentary self-efficacy to resist NSSI (mean, non-zero proportion, PAC, and PUV). The baseline-feature-based SVM and temporal-feature-based SVM were constructed using these two sets of predictors, respectively.

Figure 3 presents the ROC curves for the two SVM. Compared with the baseline-feature-based SVM, the temporal-feature-based SVM showed higher sensitivity (0.33 vs. 0.17) and negative predictive value (0.69 vs. 0.97), but showed lower specificity (0.82 vs. 0.91); the two models were comparable in accuracy (0.65) and positive predictive value (0.50). In the test set, the temporal-feature-based SVM achieved an AUC of 0.83, which surpasses both the chance level and the baseline-feature-based SVM (AUC = 0.68). These findings indicate that the temporal-feature-based model substantially outperformed the baseline-feature-based model in predicting intervention-related NSSI risk.

**Figure 3.**
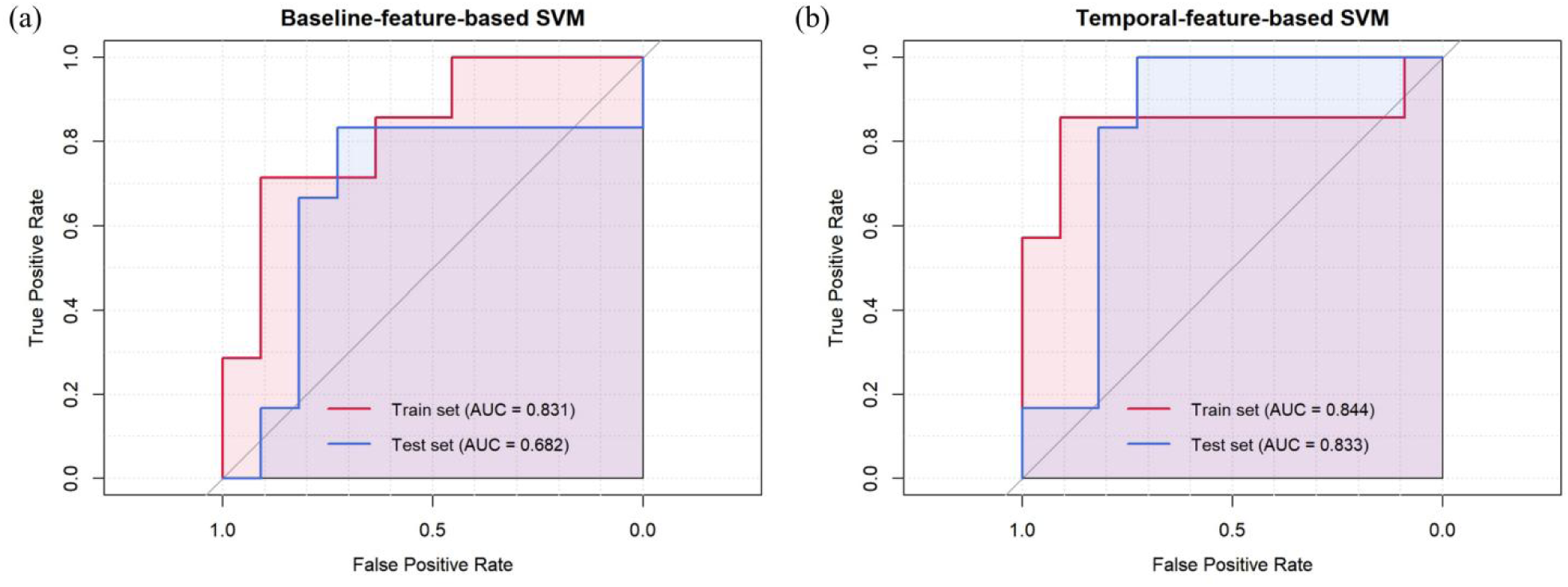
Receiver Operating Characteristic (ROC) analysis of baseline-feature-based model (a) and temporal-feature-based model (b)

## 4 Discussion

The present study employed ecological momentary assessment (EMA) and a person-centered approach to explore subtypes of NSSI ideation during inpatient care among Chinese inpatient female adolescents with mood disorders and co-occurring NSSI. Two distinct subtypes were identified, including the high-mean paroxysmal subtype and the low-mean sporadic subtype. In terms of group differences, adolescents in the high-mean paroxysmal subtype continued to show higher levels of depressive symptoms and self-harm urges than the low-mean sporadic subtype both before and after treatment, along with lower self-esteem, greater difficulties in emotion regulation, more emotional neglect experiences, and higher impulsivity. Moreover, the temporal features derived from first-week EMA outperformed baseline predictors in detecting post-discharge NSSI risk.

### 4.1 Subtypes of NSSI ideation among inpatient adolescents

Unlike previous studies that relied on retrospective data with broad time intervals, the present study employed a five-times-daily EMA sampling protocol during inpatient treatment and used temporal features as subtype indicators. Our results revealed two distinct subtypes among inpatient female adolescents with mood disorders, supporting the substantial heterogeneity of NSSI ideation in this population. Prior studies based on behavioral features have typically reported more subtypes, including three subtypes among outpatient adolescents (Lin et al., 2025) and five subtypes among Chinese inpatients with a history of suicide (Wu et al., 2022). This discrepancy in the number of identified subtypes may be due to differences in research focus. Whereas prior studies have focused on enacted features of NSSI behavior (Fox et al., 2020), the current study targets NSSI ideation, a relatively more proximal and internal psychological process. Research has documented that individuals with intense but short-lived episodes of NSSI ideation are more likely to engage in actual NSSI behaviors (Fitzpatrick et al., 2020). In that case, the assessment and subtyping of NSSI ideation in the present study may yield different results from those based on behavioral manifestations. Another possible explanation is that the participants in this study were all inpatient adolescents, who may be clinically homogeneous in terms of diagnosis and symptomatology, thereby favoring a parsimonious solution.

A critical discovery was that two subtypes were distinguished not only by overall magnitude but also across multiple temporal features. Specifically, with the exception of max change and PAC, the high-mean paroxysmal subtype showed significantly higher values on other six temporal features, suggesting a pattern characterized by sustained high levels and marked volatility in NSSI ideation. Contrary to our expectation, however, the high-mean paroxysmal subtype exhibited a significantly lower PAC than the low-mean sporadic subtype. This may be attributable to a ceiling effect: owing to the sustained high ideation in the high-mean paroxysmal subtype, consecutive changes rarely cross extreme thresholds. In contrast, the low-mean sporadic subtype, characterized by lower overall levels, was more prone to occasional sharp increases that were more readily detected as acute changes. Results from change point analyses further supported this interpretation. Despite higher PAC, the low-mean sporadic subtype displayed only a single change point, suggesting that these occasional fluctuations did not did not translate into distinct change point intervals. These findings indicate that sustained elevated states, rather than episodic acute fluctuations, may be more indicative of high risk and unfavorable treatment outcomes.

### 4.2 Between-subtype differences in treatment outcomes and individual characteristics

This study systematically compared differences in treatment outcomes and individual traits between the two subtypes, supporting the clinical value of subtyping based on temporal features. Despite significant post-intervention improvements in both subtypes, the high-mean paroxysmal subtype continued to exceed the clinical cutoffs and remained in the major depression range. Consistent with previous evidence-based research, comprehensive psychological intervention showed heterogeneous treatment responses, with a subset of adolescents continued to exhibit symptom progression and elevated future risk behaviors after intervention (Kothgassner et al., 2021). These findings underscore the importance of precisely characterize adolescents who are less responsive to standard treatment, so as to facilitate earlier and more targeted intervention.

In terms of individual characteristics, the high-mean paroxysmal subtype in the present study was characterized by more severe childhood emotional neglect, lower self-esteem, greater difficulties in emotion regulation, and higher impulsivity. These factors collectively contribute to the psychological vulnerability of this subgroup, rendering them more prone to experiencing NSSI ideation when facing negative mood or external stressors (Lee et al., 2024). Similarly, Santangelo et al. (2016) used EMA and found that female adolescents engaging in NSSI exhibited significantly lower positive affect and self-efficacy than healthy controls, and that affective instability was associated with multiple psychopathological diagnoses. Regarding impulsivity, the urgency theory posits that individuals with higher trait impulsivity may be particularly prone to impulsive actions under negative affect, as long-term consequences are outweighed by the immediate emotional relief afforded by impulsive behavior (Cyders & Smith, 2008). NSSI is also considered an impulsive behavior (Baer et al., 2018), which helps explain why individuals with higher impulsivity are prone to the more severe and fluctuating NSSI ideation. From a neurobiological perspective, the accumulation of psychological vulnerabilities may contribute to morphological changes in the anterior cingulate cortex, dorsolateral prefrontal cortex, and orbitofrontal cortex, regions that are critically involved in emotion regulation and inhibitory control (Bounoua et al., 2022). In other words, the high-mean paroxysmal subtype may represent a phenotype with neural vulnerabilities in affective regulatory circuits, heightened reactivity to both internal and external cues, and elevated trait impulsivity.

### 4.3 Predictive value of EMA-derived dynamic features for treatment response

This study revealed that the temporal-feature-based model significantly outperformed the baseline-feature-based model in predicting post-treatment NSSI urge risk, which aligns with findings among inpatient adults at high suicide risk by Wang et al. (2021). The top-10 temporal features extracted via PCA primarily encompassed the mean value, non-zero proportion and RMSSD of the three momentary assessments of NSSI ideation. Collectively, these features may reflect the stability of an individual’s NSSI ideation system during the early phase of treatment. According to dynamical systems theory, such multidimensional instability may indicate an alternative basin of attraction with low resilience (Scheffer et al., 2024). Under these circumstances, more intensive intervention is required to shift the system out of this state. This may explain why the high-mean paroxysmal subtype maintained elevated levels of clinical symptoms even after comprehensive psychological intervention.

Notably, the temporal-feature-based model in this study only incorporated EMA data collected during the first week of admission. This suggests that intensive monitoring of NSSI ideation patterns as early as one week into treatment can achieve reasonably accurate prediction of the post-treatment NSSI urge risk. Traditionally, treatment efficacy evaluation relies on pre- to post-treatment changes on standardized scales, which often requires weeks or even months to determine treatment effectiveness (Qu et al., 2023). The present findings, however, indicate that early-treatment dynamic of NSSI ideation may serve as a powerful prognostic marker, enabling clinicians to detect individuals at risk of non-response early enough to facilitate timely adjustments of treatment.

### 4.4 Limitations and implications

Several limitations are noteworthy. First, this study was conducted in a closed inpatient ward setting with a relatively small sample of female adolescents only. Although this sample composition aligns with the clinical reality that female patients constitute the vast majority of hospitalized adolescents with NSSI (Moloney et al., 2024), the small sample size and gender bias may limit the generalizability of our findings. Future research should Future research should adopt multicenter collaboration to incorporate diverse adolescent populations, such as outpatients and male adolescents. Second, the current study focused solely on NSSI ideation rather than actual NSSI behaviors. Future EMA protocols could consider investigating both ideation and behaviors simultaneously, so as to examine their heterogeneity and transition dynamics during treatment. Third, our follow-up was restricted to the inpatient period and discharge. The high-mean paroxysmal subtype retained elevated self-injury risk at discharge, indicating that this subtype may be more prone to relapse after returning to daily life (Mason et al., 2025). Given this situation, future research should increase the longitudinal follow-ups to track subtype-specific prognostic trajectories. Fourth, subtype differences were examined only in clinical symptoms and psychological traits. Integrating EMA-based dynamic features with neurophysiological measures would help characterize subtype profiles and explore the neural underpinnings of NSSI ideation dynamics.

In conclusion, this study is the first to adopt EMA-based subtyping among Chinese adolescents with mood disorders and NSSI, offering new clinical evidence for the dynamic characterization of within-group heterogeneity in this population. Theoretically, our findings expands the conceptual framework of NSSI heterogeneity. While prior studies have shown that individuals with NSSI differ on actual behavioral characteristics (Wang et al., 2021), the present study demonstrates that temporal features of NSSI ideation can also serve as valid subtyping indicators. These temporal features may capture the underlying processes related to stress reactivity and impulse control that are not accessible through retrospective assessments. In other words, heterogeneity in adolescent NSSI manifests not only in *what* is present (e.g. forms and functions) but also in *how* it unfolds over time (e.g. fluctuation patterns and propensity for sudden changes). Consisted with the integrated theoretical model of NSSI (Nock, 2009), our findings shows that both the occurrence and temporal fluctuation patterns of NSSI are influenced by distal vulnerability factors and proximal triggering factors, such as trait impulsivity and difficulties in emotion regulation. Moreover, this work preliminarily explored the feasibility of combining EMA with machine-learning in adolescent NSSI research, an approach previously used mainly for predicting suicidal behaviors (e.g., Wang et al., 2021; Wu et al., 2022). Our findings provide a methodological blueprint for using temporal features in future retrospective subtyping and prospective prediction of treatment response.

Regarding practical implications, this study offers valuable insights into matching risk profiles to interventions strategies. DBT-A, a NICE-recommended first-line treatment for adolescent NSSI, currently faces challenges of high cost and limited clinical resources. Our findings suggest that adolescents in the high-mean paroxysmal subtype, characterized by higher NSSI ideation intensity, greater fluctuation, and heightened psychological vulnerability, should be prioritized for full DBT-A intervention, especially its emotion regulation module. Targeted and comprehensive interventions that focus on NSSI ideation should be further applied to this vulnerable group. In contrast, although the low-mean sporadic subtype also exhibit clinically significant NSSI risk, their ideation is less intense and less fluctuating, and they demonstrate relatively better self-esteem and emotion regulatory capacities. For these individuals, feasible cognitive behavioral therapy (CBT) protocols targeting cognitive restructuring and problem-solving may be adequate to maintain therapeutic benefits. Such a stratified strategy can optimize clinical resource allocation, ensuring that limited therapeutic resources are preferentially channeled to adolescents with the greatest needs. In addition, this study demonstrates the feasibility of EMA in inpatient adolescents with NSSI and shows that dynamic monitoring as early as the initial phase of treatment can can provide clinically useful prognostic information. Accordingly, in clinical practice, particularly in inpatient settings, detailed monitoring of the frequency, intensity, and variability of NSSI ideation during the early admission period is warranted to enable timely identification of patients with persistently elevated and fluctuating NSSI ideation, allowing for prompt adjustments to treatment strategies.

## 5 Conclusions

The current study used intensive longitudinal data from ecological momentary assessment and identified two subtypes of NSSI ideation among Chinese inpatient female adolescents with mood disorders and co-occurring NSSI, Compared with the low-mean sporadic subtype, the high-mean paroxysmal subtype exhibited higher NSSI ideation intensity, greater short-term fluctuation, more severe clinical symptoms, and more pronounced emotion dysregulation and impulsivity. Temporal features from first-week EMA data predicted post-discharge NSSI risk with reasonable accuracy. These findings extend our understanding of the heterogeneity in NSSI ideation dynamics among adolescents with mood disorders and offer important insights for matching individual risk profiles to intervention strategies. Intensive dynamic monitoring may serve as a promising tool for clinical stratification and treatment monitoring.

## Data Availability

All data produced in the present study are available upon reasonable request to the authors.

